# Multi-class Modeling Identifies Shared Genetic Risk for Late-onset Epilepsy and Alzheimer’s Disease

**DOI:** 10.1101/2024.02.05.24302353

**Authors:** Mingzhou Fu, Thai Tran, Eleazar Eskin, Clara Lajonchere, Bogdan Pasaniuc, Daniel H. Geschwind, Keith Vossel, Timothy S Chang

## Abstract

**Background:** Previous studies have established a strong link between late-onset epilepsy (LOE) and Alzheimer’s disease (AD). However, their shared genetic risk beyond the *APOE* gene remains unclear. Our study sought to examine the shared genetic factors of AD and LOE, interpret the biological pathways involved, and evaluate how AD onset may be mediated by LOE and shared genetic risks.

**Methods:** We defined phenotypes using phecodes mapped from diagnosis codes, with patients’ records aged 60-90. A two-step Least Absolute Shrinkage and Selection Operator (LASSO) workflow was used to identify shared genetic variants based on prior AD GWAS integrated with functional genomic data. We calculated an AD-LOE shared risk score and used it as a proxy in a causal mediation analysis. We used electronic health records from an academic health center (UCLA Health) for discovery analyses and validated our findings in a multi-institutional EHR database (All of Us).

**Results:** The two-step LASSO method identified 34 shared genetic loci between AD and LOE, including the *APOE* region. These loci were mapped to 65 genes, which showed enrichment in molecular functions and pathways such as tau protein binding and lipoprotein metabolism. Individuals with high predicted shared risk scores have a higher risk of developing AD, LOE, or both in their later life compared to those with low-risk scores. LOE partially mediates the effect of AD-LOE shared genetic risk on AD (15% proportion mediated on average). Validation results from All of Us were consistent with findings from the UCLA sample.

**Conclusions:** We employed a machine learning approach to identify shared genetic risks of AD and LOE. In addition to providing substantial evidence for the significant contribution of the *APOE-TOMM40-APOC1* gene cluster to shared risk, we uncovered novel genes that may contribute. Our study is one of the first to utilize All of Us genetic data to investigate AD, and provides valuable insights into the potential common and disease-specific mechanisms underlying AD and LOE, which could have profound implications for the future of disease prevention and the development of targeted treatment strategies to combat the co-occurrence of these two diseases.

## Background

Aging is a worldwide phenomenon that significantly challenges healthcare systems and societies. As the population ages, the prevalence of age-related neurological disorders increases, including Alzheimer’s disease (AD) and late-onset epilepsy (LOE). There is increasing evidence suggesting that there may be an association between these two diseases, with people with AD being more likely to develop epilepsy and vice versa (1–7).

One potential explanation of the underlying association between AD and LOE is that shared genetic risk factors contribute to both diseases. Large genome-wide association studies (GWASs) have identified many common genetic risk factors of AD, including a major independent genetic risk factor, *APOE* (8). While there have been many GWASs conducted for AD, there is a lack of such studies for LOE, which limits the examination of shared genetic risk factors between these two conditions using cross-trait meta-analysis methods such as conjunctional false discovery rate (9) or cross-trait linkage disequilibrium score regression (LDSC) (10) to estimate the genetic correlation and shared loci between the two conditions. Moreover, the shared genetic risk factors of AD and LOE have only been identified through observational studies, with *APOE* being a well-studied example (11–13). However, the mechanisms between AD genetic risks and LOE beyond *APOE* remain unknown.

In our current study, we aim to explore the shared genetic factors of AD and LOE and interpret the potential biological pathways that link these genetic risk factors to the development of AD and LOE. The outcomes of this study may contribute to a better understanding of the disease mechanisms of AD and LOE, which would inform the development of targeted prevention and treatment strategies for these neurological disorders. Furthermore, methods used in our current study may have broader applications in other complex diseases to evaluate shared genetic risks, providing valuable insights into potential causal mechanisms and pathways.

## Methods

### Data sources

#### UCLA ATLAS Community Health Initiative

Our primary analytical sample was recruited from the biobank-linked electronic health records (EHR) of the UCLA Health System (14). The UCLA ATLAS Community Health Initiative (ATLAS) collects biosamples of participants across the UCLA Health System and associates the biosamples with the patient’s de-identified EHR. Comprehensive information about biobanking and consenting procedures can be found in previous publications (15,16). This study was considered human subject research exempt because all genetic and EHRs were de-identified (UCLA IRB# 21-000435).

#### All of Us Research Hub

We validated our findings using data from the All of Us Research Hub. The All of Us Research Hub is a centralized repository of data and tools that provides secure access to de-identified data from All of Us participants (17). We used the most recently released data (version 7), which includes 409,420 total individuals and 245,400 with whole genome sequencing.

### Sample selection

To ensure the integrity of the study and maintain a focus on longitudinal patient records, we restricted our analyses to patients with complete demographic information (age and sex) who had at least two medical encounters after the age of 60. In addition, we applied an age restriction between 60 and 90 based on the following considerations: 1) ensuring patients are old enough to have LOE (classified as onset after the age of 60); and 2) EHR data limitations: patients aged 90 or older are all marked as 90 in the UCLA de-identified EHR to maintain privacy. The final analytical sample was obtained following these selection criteria. Distributions of baseline characteristics were compared across LOE status and AD status. χ^2^ test and analysis of variance were used as appropriate to examine the homogeneity across groups.

### Phenotype definition

To define individuals with LOE or AD, we used a previously validated set of phecodes, which are groups of International Classification of Diseases, Tenth Revision (ICD-10) codes representing a specific clinical phenotype (18). The phecodes used in this study for LOE were "345" (Epilepsy, recurrent seizures, convulsions), "345.1" (Epilepsy), "345.11" (Generalized convulsive epilepsy), "345.12" (Partial epilepsy), and "345.3" (Convulsions). Individuals were considered LOE cases if they 1) had at least one encounter with any of these phecodes and 2) the age of first LOE diagnosis was after 60. For AD, we used the phecode "290.11" and defined AD cases as individuals with at least one encounter with this code.

We defined individuals as non-LOE or non-AD controls if they 1) had no diagnosis of LOE/AD and 2) had no diagnosis of phecodes within the “exclude ranges” of phecodes for each phecode mentioned above. These exclusion criteria were used to filter out controls with related conditions and to provide more accurate and reliable data for EHR case-control analysis (19). Detailed phecodes and related exclude ranges, descriptions, and mapped ICD-10 codes used in our phenotype definition are shown in **Supplementary Table 1**.

We also collected data on other health conditions that may be associated with LOE and/or AD, including hypertension (phecode "401.1"), diabetes (phecode "250.2"), and hyperlipidemia (phecode "272.1"). All ICD-phecodes mappings were performed using the R package “*PheWAS*” (20).

### Genetic data pre-processing

#### Quality control

For our UCLA ATLAS sample, we employed rigorous quality control (QC) measures and genotype imputation procedures on the genetic data to ensure the robustness and reliability of our further analyses. Details can be found elsewhere (14). After QC measures and imputation, M = 21,220,668 genotyped single nucleotide polymorphisms (SNPs) remained across N = 54,935 individuals.

#### Inferring genetic ancestry

Genetic ancestry refers to the most recent geographic ancestor of an individual’s genome, with minimal or no connection to the cultural facets of their identity (21). Genetic inferred ancestry (GIA) uses genetic data, a reference population, and inference methods to characterize individuals within a group who likely share recent geographical ancestors (22). For our UCLA ATLAS sample, we employed principal component analysis (PCA) (23) to infer a patient’s genetic ancestry using the reference panel from the 1000 Genomes Project (24). For example, we referred to “European American GIA” as individuals within the United States whose recent biological ancestors were inferred to be of European ancestry. Ancestry-specific principal components (PC) were then re-calculated using PCA to identify other population structures within each ancestry group.

#### APOE-ε4

The *APOE* gene codes for a protein that binds and transports low-density lipids and is responsible, in part, for removing cholesterol from the bloodstream. Using the two variants (rs7412 and rs429358) that define the *APOE* genotype (25), we constructed a numeric variable, “e4count”, to represent the number of ε4 alleles (0, 1, or 2) an individual carries.

### AD polygenic risk scores

A polygenic risk score (PRS) is a summary score that estimates an individual’s genetic risk for a complex disease or trait based on the collective effects of many genetic variants. It is calculated by combining information from many SNPs associated with the disease or trait of interest in GWAS (26). We selected the late-onset AD GWAS conducted by Kunkle et al. (8) for our AD PRS building based on its large sample size (21,982 cases and 41,944 controls).

We used the LDpred2 tool to build our PRSs (27). For our AD PRS without the *APOE* gene region, we removed all variants on chromosome 19 near *APOE* (from the start position of *TOMM40* − 10 kilobases to the stop position of *APOC1* + 10 kilobases) from the summary statistics. This region was removed due to the dense linkage disequilibrium (LD) block overlapping of these three genes (*TOMM40*, *APOE*, *APOC1*) in the European ancestry (28). There were M = 1,040,300 and M = 1,040,285 SNPs finally used for the building of AD PRS full and AD PRS without the *APOE* region, respectively. We also normalized all PRSs (mean = 0, standard deviation = 1) to the 1000 Genome European genetic ancestry reference population.

### Candidate SNPs identification and annotation

To identify shared genetic risk SNPs, we first identified a set of independent, genome-wide-significant, and potential causal SNPs from the AD GWAS (8). We employed the Functional Mapping and Annotation (FUMA) (29), a tool that leverages information from biological data repositories and other resources to annotate and prioritize those SNPs. We identified genomic risk loci with a P-value threshold (< 5e-8) and pre-calculated LD structure (r^2^ < 0.6) based on the European sample from the 1000 Genomes (24). Next, the mapping from SNPs to genes was obtained by performing ANNOVAR (30) using Ensembl genes (build 85). SNPs were mapped to genes through three approaches: (i) physical position on the genome, (ii) expression quantitative trait loci (eQTL) associations, and (iii) 3D chromatin interactions. Rather than GWAS P-values, we prioritized SNPs based on their functional consequences on genes. The Combined Annotation-Dependent Depletion (CADD) score (31) was used to select potential causal SNPs. We selected the SNP with at least one positional/eQTL/chromatin interaction mapped gene and with the highest CADD score within each genomic risk locus, indicating a higher likelihood that the variant is deleterious.

A total of 162 SNPs were identified as independent AD causal SNPs from the GWAS summary statistics and were subsequently used as candidate features in the AD-LOE shared genetic risk loci identification.

### Statistical analyses

#### Associations between AD genetics, AD, and LOE

We first examined AD and LOE associations in our analytic sample with logistic regression models. Then we examined the associations between AD genetic factors and AD, as well as AD genetic factors and LOE. Our primary models were adjusted for age at the last visit, sex, EHR record length, EHR record density (encounters per year), and the first four ancestry-specific PCs. Additional adjustments included health risk factors associated with AD, i.e., hypertension, diabetes, and hyperlipidemia status. Odds ratios (OR) and their 95% confidence intervals (CIs) were reported.

#### Identification of shared risk genetic loci of AD and LOE with multi-class models

A two-stage machine learning model (32) has demonstrated an improvement in prediction within multi-class modeling, surpassing the performance of individual models developed for each class. We build on this multi-class modeling paradigm using a two-step Least Absolute Shrinkage and Selection Operator (LASSO) workflow (33) to identify shared and disease-specific risk SNPs for AD and LOE (**Figure 1**). In step 1, we trained a LASSO model to predict cases with either AD or LOE using independent AD risk SNPs selected from the AD GWAS (8). Features selected by LASSO in this step were defined as the shared genetic risk loci for AD and LOE. We also defined the predicted value from this step’s model fitting as the AD-LOE risk score. In step 2, we trained another LASSO model to predict cases for specific diseases (AD or LOE) with the predicted value from step 1 as a new feature, in addition to the same features used in step 1. Features selected in this step were defined as disease-specific features.

**Figure 1.**
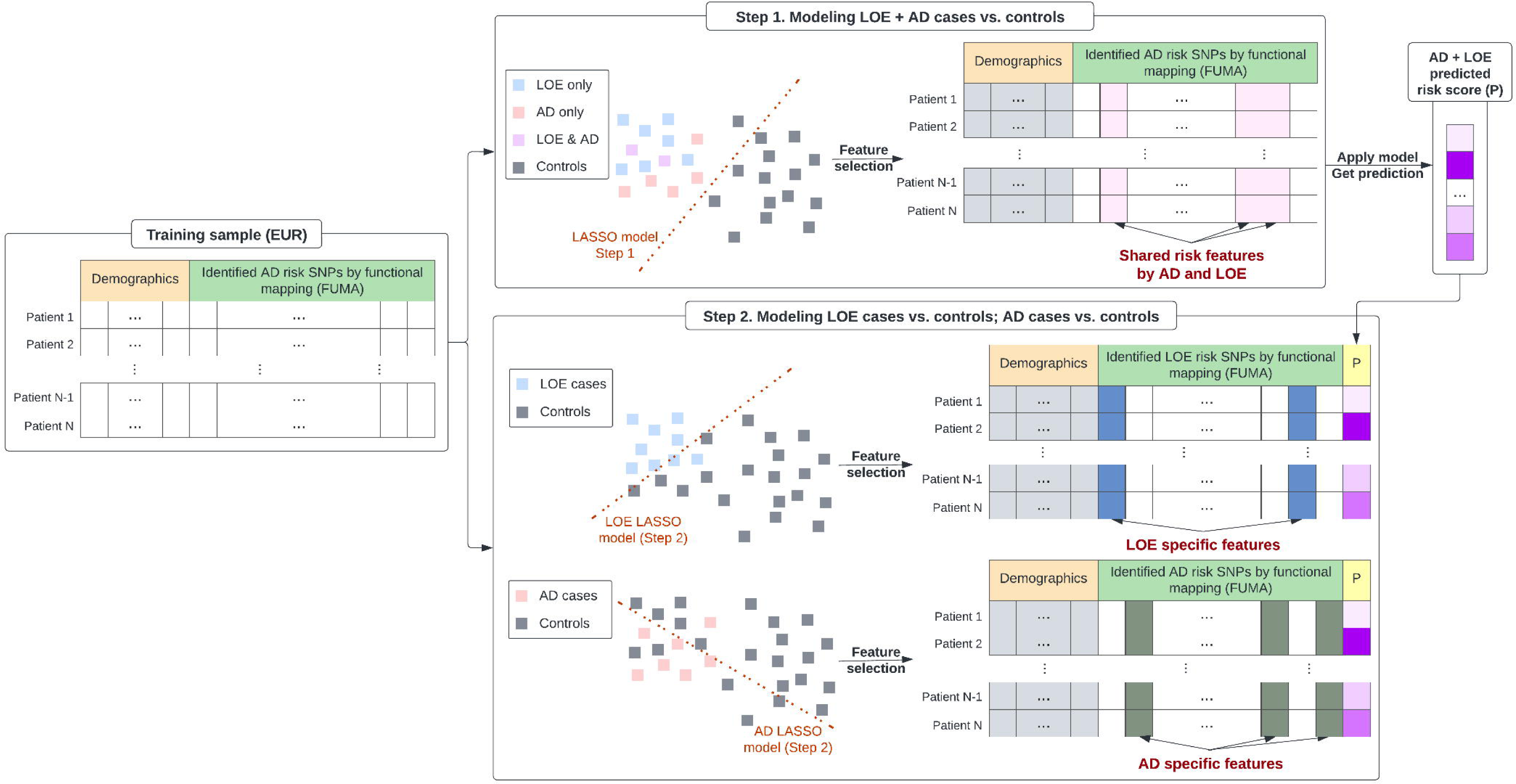
Two-step LASSO model description. In step 1, we used demographic variables (age at last visit, sex, EHR record length, EHR record density, and first four ancestry-specific principal components) and independent AD risk SNPs integrated with functional annotation from FUMA as features to train LASSO models to predict both AD + LOE as cases vs. control. Identified features from LASSO were treated as shared risk feature for these two diseases. The predicted value from step 1 (P) was used as an additional feature together with the full demographics and AD risk SNPs features in step 2. In step 2, we trained LOE cases vs. controls and AD cases vs. controls in two different LASSO models. Identified features from those models were treated as LOE-specific and AD-specific features, respectively.

For biological interpretations, we mapped identified risk SNPs from each step to genes using FUMA’s positional, eQTL, and chromatin interaction mapping (29). A set of genes of interest (e.g., mapped shared risk genes of AD and LOE) were tested against gene sets obtained from MsigDB to test for enrichment of biological functions using hypergeometric tests. Gene sets with adjusted P-value ≤ 0.05 and overlapping genes > 1 were reported and visualized with heatmaps.

#### Shared AD-LOE risk score on AD and LOE diagnosis

We further divided the predicted AD-LOE risk score into quintiles for statistical testing, allowing us to compare high-risk (top 20%) and low-risk (bottom 20%) groups. Descriptive statistics were calculated to compare various characteristics between high- and low-risk groups. Pearson’s Chi-squared or Wilcoxon rank sum test was applied appropriately to assess statistical significance.

#### Mediation analysis with shared AD-LOE risk score

We also conducted a causal mediation analysis using the predicted AD-LOE risk score (in quintiles) as the exposure, LOE as the mediator, and AD as the outcome to uncover potential mechanisms of the shared genetic risks (high vs. low risk) affecting the onset of AD through LOE. To ensure that the mediator precedes the outcome, individuals who had LOE after AD were excluded. To account for confounding effects, age at the last visit, sex, EHR record length, EHR record density, the first four ancestry-specific PCs, hypertension, diabetes, and hyperlipidemia status were adjusted. The mediation analyses were performed using the R package *’mediation’*, which offers a robust framework for investigating causal relationships with nonparametric bootstrapping (34). The mediation results were reported in terms of direct effects, indirect effects, and proportion mediated, all accompanied by 95% CI (1000 simulations).

### Sensitivity analyses

#### Model evaluation for AD or LOE prediction

Besides identifying shared risk features from step 2, we also evaluated model performances in predicting AD or LOE as a secondary analysis. We compared the performance of our two-step LASSO model to 1) benchmark model: a logistic regression model using demographic variables only; 2) AD genetic score models: logistic regression models using demographic variables and ε4 allele count/AD PRS full/AD PRS without *APOE*; 3) direct LASSO model: a LASSO model using demographic variables and independent AD risk SNPs as features for selection directly (i.e., step 2 only without step 1). A 5-fold cross-validation approach was employed, and the final results were reported on a combined hold-out testing set. Cases were upsampled to address the class imbalance issue to achieve a case-to-control ratio of 1:5, enhancing the model’s ability to accurately predict the minority class (AD or LOE cases).

The primary evaluation metric utilized was the Area Under the Precision-Recall Curve (AUPRC), which is particularly suited for imbalanced datasets (35). Additionally, the Area Under the Receiver Operating Characteristic Curve (AUC) was reported to provide a comprehensive evaluation of the model. The Matthews Correlation Coefficient (MCC) was maximized to select the optimal threshold. Other performance metrics, including F1 score, accuracy, precision, recall, and specificity, were reported based on this threshold.

#### Validations in the All of Us sample

In order to examine the generalizability of our findings from the UCLA ATLAS sample, we validated the findings using the All of Us database. We extracted a corresponding sample (European American GIA) from the All of Us Research Hub with the same criteria and employed the same approaches to define the cases and controls for AD and LOE. We also extracted the shared genetic risk loci (N = 34) identified through the step-1 LASSO model (on UCLA ATLAS sample) from the All of Us Whole Genome Sequencing (WGS) data.

It is important to note that since different SNP sets were used to infer genetic ancestry (micro-array data in UCLA ATLAS vs. WGS in All of Us), the PCA loadings calculated by the two data sources could be vastly different. Consequently, the ancestry-specific PC weights may not be directly transferable from UCLA to the All of Us sample. To address this, we retrained our model without ancestry-specific PCs on the UCLA ATLAS sample (keeping all other variables the same) and applied the updated weights to the All of Us sample (European American GIA), obtaining the predicted AD-LOE scores.

We then compared outcomes and covariates for high- and low-risk score groups for the All of Us sample, which was defined using the cutoffs from the UCLA ATLAS sample (creating quintiles and identifying the cutoffs that define the top and bottom 20%).

All analyses were performed in R statistical software (version 4.2.2) (36). We considered P-value < 0.05 for statistical significance if not specified.

## Results

### UCLA ATLAS Sample description

**Table 1** shows baseline descriptive statistics for our UCLA ATLAS sample of European American GIA. Among our primary analytic samples (N = 17,031), patients with LOE or AD have higher AD PRS (full) compared to controls. The distributions of *APOE-ε4* count and AD PRS (without *APOE*) are significantly different in AD cases and controls but do not differ in LOE cases and controls.

**Table 1.**
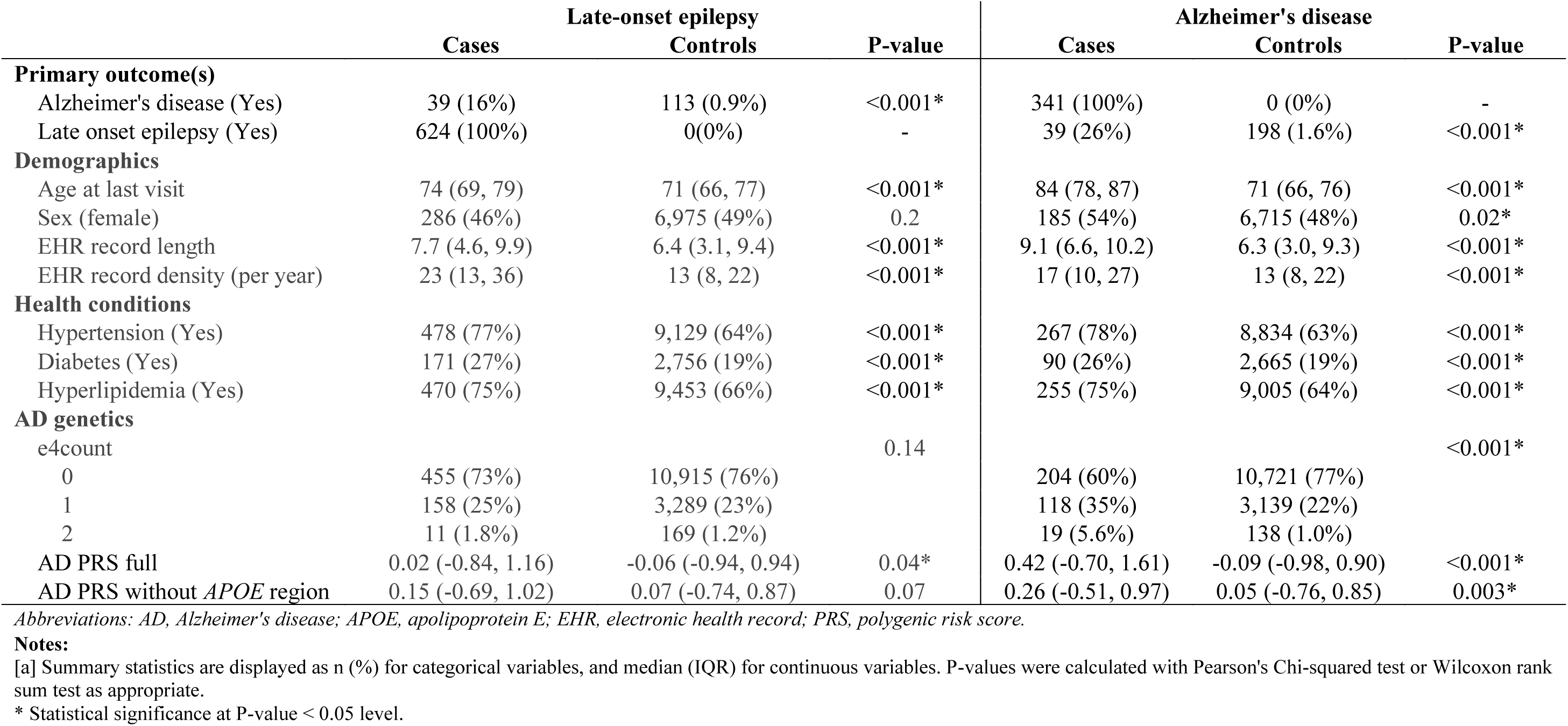
Descriptive statistics of the analytical sample by late-onset epilepsy and Alzheimer’s disease status, UCLA ATLAS patient population (N = 17,031)^a^.

|  | Late-onset epilepsy |  |  | Alzheimer's disease |  |  |
| --- | --- | --- | --- | --- | --- | --- |
|  | Cases | Controls | P-value | Cases | Controls | P-value |
| **Primary outcome(s)** |  |  |  |  |  |  |
| Alzheimer's disease (Yes) | 39 (16%) | 113 (0.9%) | <0.001* | 341 (100%) | 0 (0%) | - |
| Late onset epilepsy (Yes) | 624 (100%) | 0(0%) | - | 39 (26%) | 198 (1.6%) | <0.001* |
| **Demographics** |  |  |  |  |  |  |
| Age at last visit | 74 (69, 79) | 71 (66, 77) | <0.001* | 84 (78, 87) | 71 (66, 76) | <0.001* |
| Sex (female) | 286 (46%) | 6,975 (49%) | 0.2 | 185 (54%) | 6,715 (48%) | 0.02* |
| EHR record length | 7.7 (4.6, 9.9) | 6.4 (3.1, 9.4) | <0.001* | 9.1 (6.6, 10.2) | 6.3 (3.0, 9.3) | <0.001* |
| EHR record density (per year) | 23 (13, 36) | 13 (8, 22) | <0.001* | 17 (10, 27) | 13 (8, 22) | <0.001* |
| **Health conditions** |  |  |  |  |  |  |
| Hypertension (Yes) | 478 (77%) | 9,129 (64%) | <0.001* | 267 (78%) | 8,834 (63%) | <0.001* |
| Diabetes (Yes) | 171 (27%) | 2,756 (19%) | <0.001* | 90 (26%) | 2,665 (19%) | <0.001* |
| Hyperlipidemia (Yes) | 470 (75%) | 9,453 (66%) | <0.001* | 255 (75%) | 9,005 (64%) | <0.001* |
| **AD genetics** |  |  |  |  |  |  |
| e4count |  |  | 0.14 |  |  | <0.001* |
| 0 | 455 (73%) | 10,915 (76%) |  | 204 (60%) | 10,721 (77%) |  |
| 1 | 158 (25%) | 3,289 (23%) |  | 118 (35%) | 3,139 (22%) |  |
| 2 | 11 (1.8%) | 169 (1.2%) |  | 19 (5.6%) | 138 (1.0%) |  |
| AD PRS full | 0.02 (-0.84, 1.16) | -0.06 (-0.94, 0.94) | 0.04* | 0.42 (-0.70, 1.61) | -0.09 (-0.98, 0.90) | <0.001* |
| AD PRS without *APOE* region | 0.15 (-0.69, 1.02) | 0.07 (-0.74, 0.87) | 0.07 | 0.26 (-0.51, 0.97) | 0.05 (-0.76, 0.85) | 0.003* |

Abbreviations: AD, Alzheimer's disease; APOE, apolipoprotein E; EHR, electronic health record; PRS, polygenic risk score.
**Notes:**
[a] Summary statistics are displayed as n (%) for categorical variables, and median (IQR) for continuous variables. P-values were calculated with Pearson's Chi-squared test or Wilcoxon rank sum test as appropriate.
\* Statistical significance at P-value < 0.05 level.

### Associations between AD genetics, AD phenotype, and LOE

In the UCLA EUR sample, after adjusting for age at the last visit, sex, EHR record length, EHR record density, and ancestry-specific PCs, *APOE-ε4* count (OR = 2.74, 95%CI: 2.22, 3.36), AD PRS (full) (OR = 1.29, 95%CI: 1.19, 1.39), and AD PRS (without *APOE*) (OR = 1.13, 95%CI: 1.02, 1.25) are all positively associated with AD, while only *APOE-ε4* count (OR = 1.21, 95%CI: 1.02, 1.43) is positively associated with LOE (**Table 2**). We also observed a strong positive association between AD and LOE (OR = 17.72, 95%CI: 11.24, 27.58). Association results are similar with further adjustment of health conditions, including hypertension, diabetes, and hyperlipidemia status at the last visit.

**Table 2.** Associations between AD phenotype, AD genetics, and late-onset epilepsy status, UCLA.

| Exposure | Covariates demographics<sup>a</sup> |  | Covariates: demographics + health conditions<sup>b</sup> |  |
| --- | --- | --- | --- | --- |
|  | OR | 95% CI | OR | 95% CI |
| **Outcome: Alzheimer's disease (N = 14,339)** |  |  |  |  |
| LOE status | 17.72 | (11.24, 27.58) | 17.57 | (11.13, 27.41) |
| e4count | 2.74 | (2.22, 3.36) | 2.77 | (2.25, 3.41) |
| AD PRS full | 1.29 | (1.19, 1.39) | 1.29 | (1.20, 1.39) |
| AD PRS without *APOE* | 1.13 | (1.02, 1.25) | 1.13 | (1.02, 1.25) |
| **Outcome: late-onset epilepsy (N = 14,997)** |  |  |  |  |
| AD status | 16.78 | (10.80, 25.74) | 16.61 | (10.68, 25.49) |
| e4count | 1.21 | (1.02, 1.43) | 1.22 | (1.03, 1.44) |
| AD PRS full | 1.06 | (1.00, 1.12) | 1.06 | (1.00, 1.12) |
| AD PRS without *APOE* | 1.06 | (0.98, 1.14) | 1.06 | (0.98, 1.14) |

Abbreviations: AD, Alzheimer's disease; APOE, apolipoprotein E; CI, confidence interval; EHR, electronic health record; OR, odds ratio; PRS, polygenic risk score.
**Notes:**
[a] Demographics adjustments include age at last visit, sex, EHR record length, EHR record density, and first four ancestry-specific principal components.
[b] Health conditions adjustments include hypertension, diabetes, and hyperlipidemia status.

### Shared genetic risks of AD and LOE with multi-class models

We leverage the multi-class machine learning paradigm using a two-step approach in order to identify shared and disease-specific genetic risks for AD and LOE. We identified 34 SNPs as the shared genetic risk factors of AD and LOE from the step-1 LASSO model, which uses AD or LOE as the outcome (**Figure 2A**). Many risk SNPs are located near *19q13.32*. Among those risk SNPs, chr19:45406538:I, rs157580, and rs1064725 are the top three SNPs having the highest coefficient magnitude. Risk SNPs located outside the *19q13.32* region, such as rs1286162 (*11q12*) and chr6:47442154:D (*6p12*), also hold substantial relevance in contributing to the shared genetic risk factors of AD and LOE, which reinforces the complexity of the genetic interplay between these two disorders.

**Figure 2.**
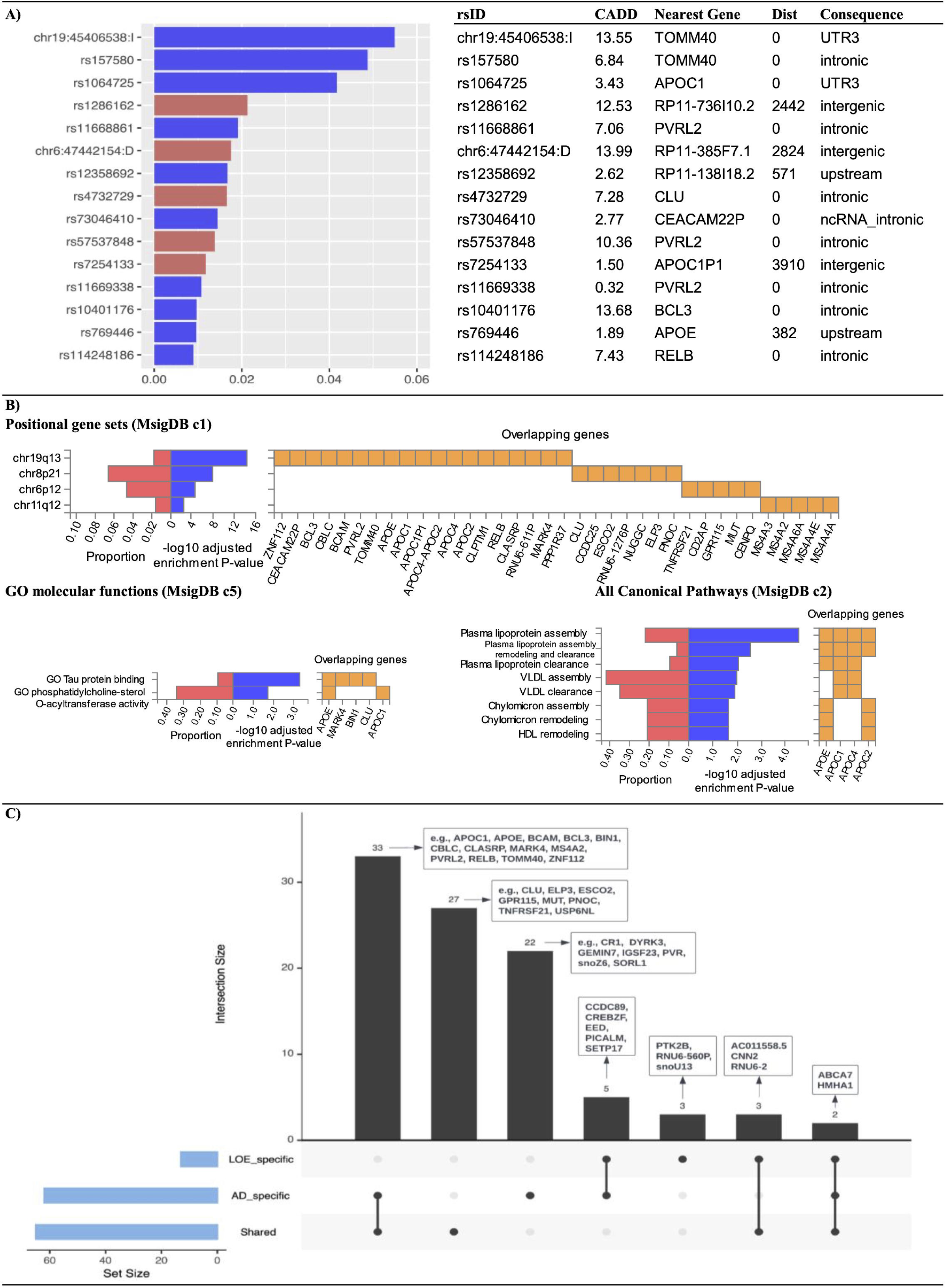
Shared risk genetic loci, mapped genes and enrichment test results from the two-step LASSO models. A) Standardized coefficient magnitude of shared risk features (top 15) identified from Step 1: Alzheimer’s disease or late-onset epilepsy as the outcome. Colors in the bar plot represent direction of effects (red for ’+’, blue for ’-’). B) Enrichment test results for shared risk genes of Alzheimer’s disease and late-onset epilepsy. The set of genes were mapped from shared risk genetic loci from the step 1. C) Summary of genes mapped from genetic loci of risk features in the two-step LASSO models.

Furthermore, in the step-2 LASSO model, we selected N = 35 risk SNPs for AD, of which 24 were not part of the shared genetic risk loci set, and 3 risk SNPs for LOE, with 2 not belonging to the shared genetic risk loci set, as disease-specific risk SNPs. Notably, rs10422350, positionally mapped to the *snoZ6* gene, and rs618679, positionally mapped to the *PICALM* gene, were identified as AD-specific risk SNPs. rs73223431 and rs567075, positionally mapped to the *PTK2B* and *snoU13* genes respectively, were identified as LOE-specific risk SNPs.

The identified risk SNPs (shared and disease-specific) were mapped to 95 genes (**Figure 2B**). We identified the *APOE* region *19q13* as an essential genomic risk locus among all mapped AD-LOE shared risk genes (N = 65). However, we also observed an enrichment at other chromosome positions, including *8p21*, *6p12*, and *11q12*. These shared risk genes also showed enrichment in two gene ontology molecular functions, including tau protein binding and phosphatidylcholine-sterol O-acyltransferase activity, and eight canonical pathways, such as plasma lipoprotein assembly, remodeling, and clearance.

We also examined the overlapping and distinct genes between shared, AD-specific, and LOE-specific gene sets (**Figure 2C**). For example, the well-known AD risk genes *APOE*, *TOMM40*, and *APOC1* were identified by shared AD-LOE risk and AD-specific risk gene sets. Additionally, *CR1*, *DYRK2*, and *snoZ6* were exclusively identified as AD-specific genes, while *PTK2B*, *RNU6-560P*, and *snoU13* were uniquely categorized as LOE-specific genes.

Importantly, two genes, *ABCA7* and *HMHA1*, were found in all three gene sets, underscoring their potential role in the shared and distinct genetic mechanisms underlying AD and LOE. The complete list of risk SNPs and their mapping information can be found in **Supplementary Table 2-5**.

### Shared AD-LOE risk score

In the UCLA ATLAS European GIA sample, individuals with high shared risk scores (top 20% of the population) compared to those with low-risk scores (bottom 20% of the population) have a higher risk of developing AD (8.2% vs. 0.3%, p-value < 0.001) or LOE (9% vs. 1%, p-value < 0.001), and are more likely to develop both conditions in their later life (both: 1.2% vs. 0%, p-value < 0.001) (**Table 3**).

**Table 3.** Descriptive statistics by predicted risk score from step-1 model (either Alzheimer’s disease or late-onset epilepsy prediction), high vs.low predicted risk^a^.

|  | UCLA ATLAS sample<sup>b</sup> |  |  | All of Us sample<sup>c</sup> |  |  |
| --- | --- | --- | --- | --- | --- | --- |
|  | High<br>N = 3,406 | Low<br>N = 3,407 | P-value | High<br>N = 3,099 | Low<br>N = 18,765 | P-value |
| **Primary outcome(s)** |  |  |  |  |  |  |
| Alzheimer's disease (Yes) | 204 (8.2%) | 8 (0.3%) | <0.001* | 50 (2.0%) | 54 (0.3%) | <0.001* |
| Age at Alzheimer's diagnosis | 82.3 (78.0, 85.9) | 64.0 (60.6, 69.2) | <0.001* | 78 (75, 83) | 66 (62, 70) | <0.001* |
| Late onset epilepsy (Yes) | 249 (9.0%) | 30 (1.0%) | <0.001* | 152 (5.7%) | 394 (2.3%) | <0.001* |
| Age at LOE diagnosis | 74 (69, 79) | 63 (61, 65) | <0.001* | 74.1 (67.7, 78.3) | 63.7 (61.6, 67.1) | <0.001* |
| Condition |  |  | <0.001* |  |  | <0.001* |
| both | 25 (1.2%) | 0 (0%) |  | 5 (0.2%) | 7 (<0.1%) |  |
| AD only | 179 (8.3%) | 8 (0.3%) |  | 45 (2.1%) | 47 (0.3%) |  |
| LOE only | 224 (10%) | 30 (1.0%) |  | 147 (6.7%) | 387 (2.4%) |  |
| none | 1,719 (80%) | 2,884 (99%) |  | 1,991 (91%) | 15,541 (97%) |  |
| **Demographics** |  |  |  |  |  |  |
| Age at last visit | 80 (76, 85) | 64 (62, 66) | <0.001* | 79 (74, 83) | 66 (63, 70) | <0.001* |
| Sex (female) | 1,749 (51%) | 1,537 (45%) | <0.001* | 1,709 (55%) | 10,787 (57%) | 0.02* |
| EHR record length | 10.0 (8.9, 13.9) | 2.3 (1.2, 3.8) | <0.001* | 11.0 (6.1, 14.8) | 3.3 (1.6, 5.2) | <0.001* |
| EHR record density (per year) | 20 (11, 32) | 11 (7, 17) | <0.001* | 13 (7, 31) | 7 (4, 12) | <0.001* |
| **Health conditions** |  |  |  |  |  |  |
| Hypertension (Yes) | 2,786 (82%) | 1,661 (49%) | <0.001* | 2,301 (74%) | 9,032 (48%) | <0.001* |
| Diabetes (Yes) | 939 (28%) | 450 (13%) | <0.001* | 726 (23%) | 3,129 (17%) | <0.001* |
| Hyperlipidemia (Yes) | 2,720 (80%) | 1,704 (50%) | <0.001* | 2,120 (68%) | 8,019 (43%) | <0.001* |

Abbreviations: AD, Alzheimer's disease; APOE, apolipoprotein E; EHR, electronic health record; LASSO, Least Absolute Shrinkage and Selection Operator; LOE, late-onset epilepsy; PRS, polygenic risk score.
**Notes:**
[a] Summary statistics are displayed as n (%) for categorical variables, and median (IQR) for continuous variables. P-values were calculated with Pearson's Chi-squared test or Wilcoxon rank sum test as appropriate.
[b] High/low predicted risk was defined as top/bottom 20% in predicted risk score from the two-step LASSO model, step 1: either Alzheimer's disease or late-onset epilepsy prediction.
[c] The predicted risk scores and high/low risk for the All of Us sample were calculated and defined based on the model and corresponding threshold from the training sample (UCLA ATLAS).
\* Statistical significance at P-value < 0.05 level.

After filtering out patients missing in either AD or LOE and who had LOE after AD, there were 12,290 UCLA ATLAS European GIA individuals used in downstream mediation analyses (**Table 4**). All of the assumptions for mediation analysis were met (**Supplementary Table 6**).

**Table 4.**
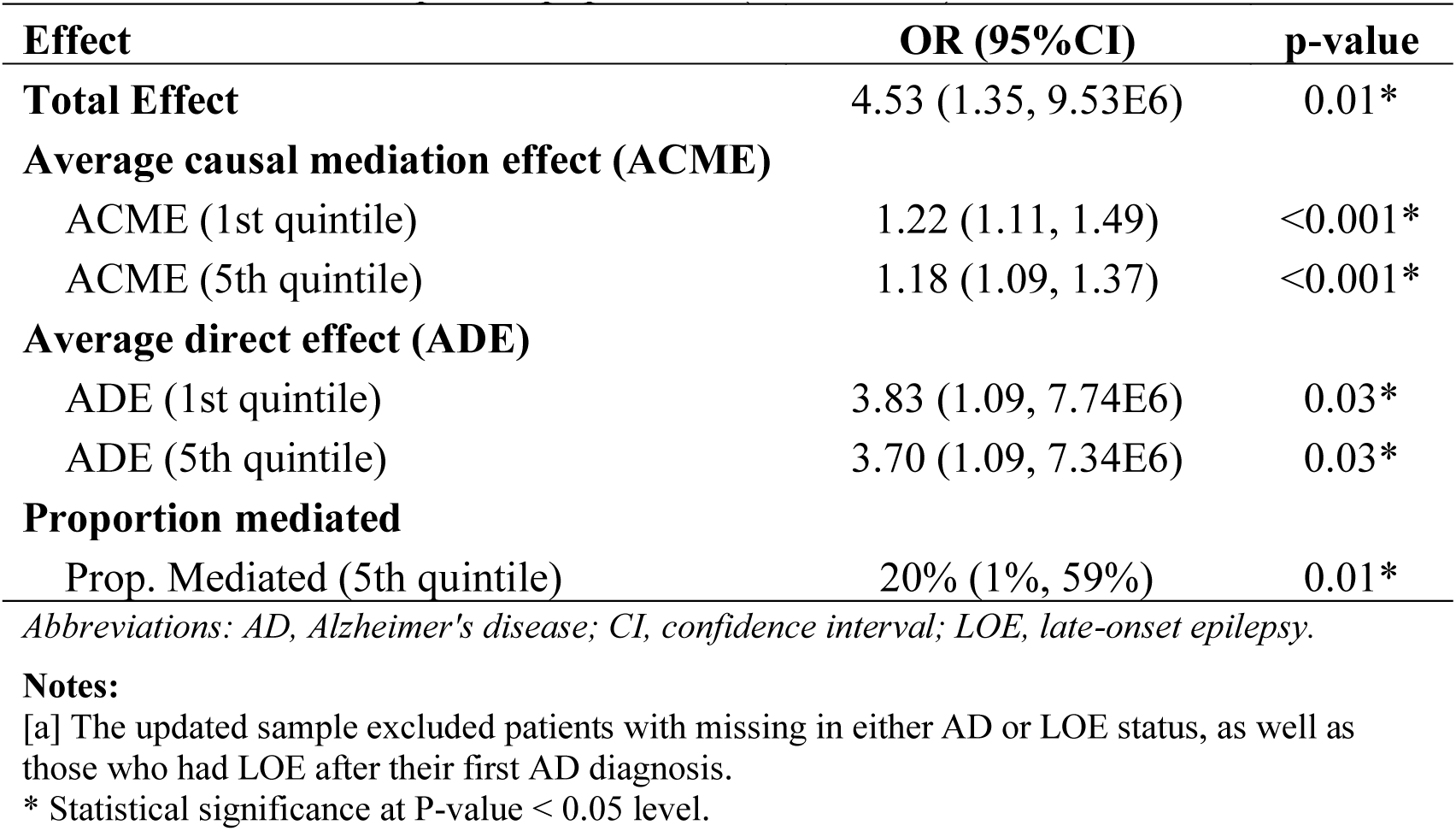
Mediation results of AD-LOE predicted risk scores, LOE, and AD status, UCLA ATLAS patient population (N = 12,290)^a^.

The total effect of the shared AD-LOE risk score on AD was significant (effect estimate = 0.014, 95%CI: 0.003, 0.02). In conjunction with the lower average direct effect (ADE) compared to the total effect, the significant average causal mediation effect (ACME) (effect estimate = 0.002, 95%CI: 0.001, 0.004) indicates that partial mediation exists. Interestingly, the mediation effect of LOE is greater for individuals with high AD-LOE genetic risk score (proportion mediated: 23% vs. 7.7%).

### Sensitivity analyses

#### Multi-class Model performance in AD or LOE prediction

Our two-step LASSO model can not only find the shared risk factors between AD and LOE but also help predict AD or LOE onset. In the comparisons among benchmark, AD genetic scores, and machine learning models, the two-step LASSO model performs the best in AD and LOE prediction. Specifically, the two-step LASSO model improves 26% of the AUPRC in AD prediction and 23% in LOE prediction compared to the benchmark model with demographics only (AD: 0.1667 to 0.2097; LOE: 0.0688 to 0.0849). The two-step LASSO model also outperformed the AD genetic score models (e4count, AD PRS (full), and AD PRS (without *APOE*)) and performed slightly better than the direct LASSO model (**Supplementary Table 7**).

#### Validation of AD-LOE shared genetic risk score in the All of Us sample

We first compared the distributions of the All of Us validation sample to our discovery UCLA ATLAS sample (**Supplementary Table 8**). Patients from these two samples differ in all variables except for diabetes status. The baseline descriptive statistics of variables by AD and LOE in the All of Us sample are shown in **Supplementary Table 9**. Similar to findings from the UCLA sample, we observed a higher proportion of AD cases in LOE cases and vice versa.

We calculated the predicted shared AD-LOE risk score for the All of Us sample by fitting the step-1 model (trained on the UCLA ATLAS sample). Consistent with our findings from the UCLA EUR sample, individuals characterized by high shared risk scores, as opposed to those with low-risk scores, exhibit a higher risk of developing AD (2% vs. 0.3%, p-value < 0.001) or LOE (5.7% vs. 2.3%, p-value < 0.001). Moreover, these individuals also have a higher likelihood of developing both conditions in their later life (0.2% vs. <0.1%, p-value < 0.001) (**Table 3**).

## Discussion

The shared genetic risks of AD and LOE remain largely unexplored, particularly beyond the well-known *APOE* gene. To bridge this knowledge gap, our current study employs a machine learning approach to identify shared genetic risks of AD and LOE within the European American GIA population. Utilizing a two-step LASSO method, we identified 34 shared risk SNPs between AD and LOE, which were further mapped to genes and studied to explore their downstream biological functions. We identified the *APOE* region as an important genomic risk locus for AD and LOE, along with other important chromosome positions, including *8p21*, *6p12*, and *11q12.* These shared risk genes showed enrichment in molecular functions and pathways such as tau protein binding, phosphatidylcholine-sterol O-acyltransferase activity, and plasma lipoprotein assembly, remodeling, and clearance. Validations on the All of Us database enhanced the reliability and generalizability of our findings.

Our results align with and build upon previous studies (11,12,37), corroborating the notion that not only the *APOE* region plays a significant role in the shared genetic risks underlying AD and LOE. We also uncovered novel genes that may contribute, such as *CLU*, located on chromosome 8p21, *TNFRSF21* on chromosome 6p12, and *MS4A* on chromosome 11q12. It is of particular interest that we identified two genes, *ABCA7* and *HMHA1*, during our examination of overlapping genes within shared, AD-specific, and LOE-specific gene sets, as depicted in **Figure 2C**. The ATP-binding cassette, sub-family A, member 7 (*ABCA7*) gene, situated between the *CNN2* and *HMHA1* genes, has been associated with the regulation of lipid metabolism and phagocytosis (38). Both in vitro and in vivo studies have demonstrated that genetic and epigenetic markers of *ABCA7* exhibit significant correlations with β-amyloid deposition and brain morphology (39,40). Our findings suggest that *ABCA7* and *HMHA1* may play a crucial role not only in the pathogenesis of AD but also in that of LOE. These findings expand the current understanding of the shared genetic landscape between AD and LOE, and warrant further investigation into their underlying mechanisms, their influence on disease onset, and potential applications in treatment strategies.

Enrichment analysis of the identified genes enhances our understanding of the pathogenesis underlying the interconnection between these two diseases. Our analysis revealed a notable enrichment in tau protein binding with identified genes such as *APOE, MARK4, BIN1, CLU*, and *APOC1*, as represented in **Figure 2B**. Tau protein, a critical microtubule-associated protein, has been implicated in the pathophysiology of epilepsy and AD, signifying its potential as a therapeutic target (41). Remarkably, tau reduction has been identified as a viable therapeutic strategy for AD and other related disorders characterized by epileptic activity. In a mouse model of AD, genetic reduction of tau levels was found to prevent learning and memory disruption, epileptic activity, and other AD-related deficits (42). Furthermore, biomarkers associated with tau, such as total tau and p-tau levels in cerebrospinal fluid (41,43,44), hold promise for predicting the incidence of LOE and AD, demonstrating their potential as valuable tools for disease prognosis. This evidence and our results suggest that tau-mediated mechanisms may provide a novel opportunity for the development of disease-modifying therapies for both LOE and AD.

Lastly, we conducted secondary analyses to explore the utility of the predicted value derived from the step-1 LASSO model. The predicted score served as a shared risk score for AD and LOE, demonstrating its versatility in various applications. Our findings showcased the shared risk score’s capability to 1) effectively stratify patients into distinct AD-LOE risk groups, allowing for better risk assessment and management; 2) predict the onset of AD and/or LOE diseases in later life, highlighting its potential as a valuable prognostic tool; and 3) examine the causal mediation of shared genetic risks between AD and LOE, providing insight into the complex interplay between these two factors and the potential consequences of their shared genetic underpinnings.

There are several notable strengths in our study. First, by leveraging advanced machine learning techniques, we were able to identify both well-established and novel genes associated with the shared risks of AD and LOE. Our machine learning approach does not require GWAS summary statistics for both diseases, making it a versatile method that can be applied to investigate shared genetic risks of other diseases, particularly those without extensive GWAS summary statistics, such as LOE. Second, the validation of our model on the All of Us database enhanced the robustness and generalizability of our findings, increasing confidence in the results and their potential implications. Finally, our investigation into the utility of the predicted risk score underscores its potential applications in various aspects of disease prevention, diagnosis, and management, fostering a more comprehensive understanding of the shared genetic landscape between AD and LOE.

While our study provides valuable insights into the shared genetic risks of AD and LOE, there are limitations worth noting. First, our study is applicable to individuals of European American GIA only. Due to the small sample sizes of other ancestry groups, we did not perform modeling for these populations. As a result, the generalizability of our findings to other ancestries is limited. Moreover, participant samples in our study were selected based on ICD-10 diagnosis codes retrieved from EHRs, which could subject to misclassifications. Nonetheless, given the substantial size of our cohort, sporadic misclassifications are unlikely to significantly impact the overarching results. To address these issues, future studies should consider training with larger sample sizes, collecting data from multiple institutions, and focusing on underrepresented populations to enhance the generalizability of findings across diverse populations.

## Conclusions

In conclusion, our study aimed to elucidate the shared genetic risks of AD and LOE in the European American GIA population by deploying a two-step LASSO method. Our study contributes to a deeper understanding of the molecular mechanisms that govern the complex interplay between neurodegenerative diseases. Moreover, we demonstrated the utility of the predicted shared-risk score in stratifying patients into distinct AD-LOE risk groups, predicting disease onset later in life, and examining causal mediation of shared genetic risks. By addressing the study’s limitations and building upon its strengths, researchers can continue to uncover the complex relationships between AD and LOE, ultimately contributing to a more comprehensive understanding of their shared genetic underpinnings.

## Data Availability

The datasets generated and/or analyzed during the current study are not publicly available because individual electronic health record data are not publicly available due to patient confidentiality and security concerns, but are available from the corresponding author (Timothy S Chang) on reasonable request.

## List of abbreviations

ACME: Average Causal Mediation Effect
AD: Alzheimer’s Disease
ADE: Average Direct Effect
APOE: Apolipoprotein E
AUC: Area Under the Receiver Operating Characteristic Curve
AUPRC: Area Under the Precision-Recall Curve
CADD: Combined Annotation-Dependent Depletion
CI: Confidence Interval
EHR: Electronic Health Record
eQTL: Expression Quantitative Trait Loci
GIA: Genetic Inferred Ancestry
GWAS: Genome-Wide Association Study
ICD-10: International Classification of Diseases, Tenth Revision
LASSO: Least Absolute Shrinkage and Selection Operator
LD: Linkage Disequilibrium
LOE: Late-Onset Epilepsy
MCC: Matthews Correlation Coefficient
OR: Odds Ratio
PC: Principal Components
PCA: Principal Component Analysis
PRS: Polygenic Risk Score
QC: Quality Control
SNP: Single Nucleotide Polymorphisms

## Declarations

### Ethics approval and consent to participate

This study was considered human subject research exempt because all genetic and EHRs were de-identified (UCLA IRB# 21-000435).

### Availability of data and materials

The datasets generated and/or analyzed during the current study are not publicly available because individual electronic health record data are not publicly available due to patient confidentiality and security concerns, but are available from the corresponding author (Timothy S Chang) on reasonable request. Code is available on GitHub: https://github.com/TSChang-Lab/AD_LOE_shared_genetics.

### Authors’ contributions

MF, TT, KV and TSC contributed to conception and design of the study. MF and TT performed the statistical analysis. MF wrote the first draft of the manuscript. TT wrote sections of the manuscript. All authors contributed to manuscript revision, read, and approved the submitted version.

## Acknowledgements

We gratefully acknowledge the resources provided by the Institute for Precision Health (IPH) and participating UCLA ATLAS Community Health Initiative patients. The UCLA ATLAS Community Health Initiative in collaboration with UCLA ATLAS Precision Health Biobank, is a program of IPH, which directs and supports the biobanking and genotyping of biospecimen samples from participating UCLA patients in collaboration with the David Geffen School of Medicine, UCLA CTSI and UCLA Health. We would also like to acknowledge all participants and researchers at the All of Us program. The All of Us Research Program is supported by the National Institutes of Health, Office of the Director: Regional Medical Centers: 1 OT2 OD026549; 1 OT2 OD026554; 1 OT2 OD026557; 1 OT2 OD026556; 1 OT2 OD026550; 1 OT2 OD 026552; 1 OT2 OD026553; 1 OT2 OD026548; 1 OT2 OD026551; 1 OT2 OD026555; IAA #: AOD 16037; Federally Qualified Health Centers: HHSN 263201600085U; Data and Research Center: 5 U2C OD023196; Biobank: 1 U24 OD023121; The Participant Center: U24 OD023176; Participant Technology Systems Center: 1 U24 OD023163; Communications and Engagement: 3 OT2 OD023205; 3 OT2 OD023206; and Community Partners: 1 OT2 OD025277; 3 OT2 OD025315; 1 OT2 OD025337; 1 OT2 OD025276.

